# Systematic review of high-dose amikacin regimens for the treatment of Gram-negative infections based on EUCAST dosing recommendations

**DOI:** 10.1101/2022.05.22.22275426

**Authors:** Kevin J. Frost, Ryan A. Hamilton, Stephen Hughes, Conor Jamieson, Paul Rafferty, Oliver Troise, Abi Jenkins

## Abstract

**Background:** Amikacin is an aminoglycoside with activity against Gram negative pathogens. Updated EUCAST amikacin breakpoints for Enterobacterales and Pseudomonas aeruginosa included revised dosing recommendations of 25-30mg/kg to achieve key pharmacokinetic/pharmacodynamic parameters, higher than recommended in the British National Formulary. We undertook a literature review to report preferred dosing regimens, monitoring and toxicities associated with the use of amikacin at doses ≥20mg/kg/day.

**Methods:** This literature search was conducted in electronic databases for any study reporting adult participants treated with amikacin at doses ≥20mg/kg/day. Data were extracted for pharmacokinetic parameters and clinical outcomes, while papers were assessed for bias using the ROBINS-I tool.

**Results:** Nine papers were identified and included, eight of which were observational studies; assessment of bias showed substantial flaws. Dosing regimens ranged from 25-30mg/kg/day. Six studies adjusted the dose in obesity when participants BMI ≥30 kg/m2. Target peak serum concentrations ranged from 60mg/L-80mg/L and 59.6-81.8% of patients achieved these targets. Two studies reported the impact of high dose amikacin on renal function. No studies reporting auditory or vestibular toxicity were identified.

**Conclusions:** Dosing amikacin at 25-30mg/kg achieved peak concentration targets in the majority of patients, but there was no information on clinical outcomes. There is little information about the impact on renal function or ototoxicity; caution with use of high dose regimens in older patients for prolonged periods is recommended. Given the paucity of information, there is a need for a consensus guideline for high dose amikacin or a prospective study.

**What is already known on this topic:** Amikacin is receiving increased interest as an antibiotic option for multidrug resistant organisms

Amikacin and other aminoglycosides require therapeutic drug monitoring to minimise the risk of nephrotoxicity

Increasing prevalence of antimicrobial resistance in key pathogens has led to changes to susceptibility breakpoints and theoretical dosing recommendations in European-wide guidelines, including a recommendation for high-dose amikacin for certain pathogens

**What this study adds:** The current literature reporting data and outcomes with high-dose amikacin regimens has a high degree of bias and is confounded by poor study design and as a result there in insufficient evidence base to provide guidance on how to manage high-dose amikacin.

Appropriate dosing weight for obese patients, adjustment for renal impairment, monitoring interval, potential toxicity and key PK/PD targets to guide treatment with high-dose amikacin regimens remain poorly defined in the current literature.

**How this study might affect research, practice or policy:** Further evidence and/or consensus guidelines based on expert judgement are required to ensure patients can receive optimal therapy when amikacin is the treatment of choice.

## Introduction

Aminoglycosides are widely used antibiotics for the treatment of Gram-negative infections and the increasing prevalence of multi-drug resistant Gram-negative organisms has renewed interest in their use [1]. Numerous resistance mechanisms to aminoglycosides exist, including the increasingly prevalent aminoglycoside-modifying enzymes (AMEs) but also 16S rRNA methyltransferases (RMTases) [2]. Amikacin is less susceptible to many of the common aminoglycoside modifying enzymes (AMEs) in Gram-negative pathogens [3], therefore it provides a valuable treatment option in patients or populations with gentamicin resistance.

Amikacin is licensed, and traditionally used, at a dose of 15 mg/kg once daily or 7.5 mg/kg twice daily [4]. Additionally, the manufacturer cautions against exceeding a single dose of 1.5 g/day and a total course of more than 15 g. There is renewed interest in high-dose amikacin regimens, based on increasing understanding of the pharmacokinetic/pharmacodynamic factors associated with target attainment and maximal killing, and limitation of mutant selection due to suboptimal dosing. Thus, understanding how to optimise dosing of amikacin to balance the treatment efficacy with known treatment toxicities (for example nephrotoxicity and ototoxicity) is increasingly important.

Recent EUCAST changes to antimicrobial susceptibility breakpoints [5] address laboratory-based technical uncertainty and concerns about achievable drug exposure *in vivo*. Through these changes, susceptibility cut-offs for antimicrobial wild type distributions of microorganisms (or epidemiologic cut-off values) are introduced with a breakpoint of 8 mg/L and 16 mg/L for Enterobacterales and Pseudomonas sp., respectively [6]. This introduces discordance with current practice and the known optimum pharmacokinetic/pharmacodynamic indices for aminoglycoside dosing. To obtain the desired bactericidal activity with amikacin (1-log reduction in growth), a pharmacodynamics target attainment of C_max_/MIC of ≥8 is proposed [7,8]. This is widely disputed, and many propose an AUC/MIC ratio (≥80-90) as a more robust alternative [5,9]. EUCAST dosing methodology is based on the latter, with Monte Carlo simulations presented with their revised breakpoints [5]. Irrespective of the PK/PD parameter used, pathogens with high MIC value (4-16 mg/L) risk sub-therapeutic dosing when amikacin is used at its current UK licensed dosing of 15 mg/kg once daily [4]. A revised dosing of 25-30 mg/kg once daily has been recommended by EUCAST to address this risk. However, there is no supporting information on how to dose and monitor patients appropriately at this higher dosing range and it presents a challenge to translate this recommendation into routine clinical practice. A previous review of standard amikacin dosing and monitoring was unable to provide any recommendations for routine dosing regimens [10].

The objectives of this review were to identify evidence for high-dose amikacin therapeutic regimens and to determine drug exposures that are related to adverse events and toxicity.

## Method

This systematic review and meta-analysis was registered in the International prospective register of systematic reviews (PROSPERO) with the reference CRD42021250022, and it was conducted following the PRISMA 2020 statement criteria [11].

This systematic review investigates therapeutic drug monitoring (TDM), dose adjustment and toxicities of high-dose amikacin. Initial plans were to update the literature search undertaken by Jenkins *et al* [10] with the addition of amikacin doses ≥20 mg/kg/day as a sub-group. Papers reporting doses ≥20mg/kg/day amikacin were searched to ensure studies investigating doses higher than the UK standard dose were captured. Following full text review no papers were identified that met the inclusion criteria. Consequently, a new protocol was developed which broadened the range of acceptable papers. Inclusion criteria comprised adults with infections treated with amikacin at doses ≥20 mg/kg/day in randomized control trials, controlled clinical trials, interrupted time series, controlled before and after studies and observational studies (see Supplementary Information – Literature Review Protocol).

Literature searches were conducted in July 2021 and updated in October 2021 using Healthcare Database Advanced Search tool and included the databases Medline, EMBASE, CINAHL, the Cochrane Central Register of Controlled Trials and Google Scholar. Reference lists of all included papers were also searched for potential papers of interest.

Papers were reviewed by title and abstract for the initial triage and then in full text on each occasion by two authors *(AJ, CJ, RH, SH, PR and OT)* for inclusion. Any disagreements were resolved by review by a third author (*KF)*. Studies were limited to those that could be accessed in full-text and were published in English.

Two authors independently assessed the risk of bias for each paper using the ROBINS-I tool [12]. Each study was assessed for bias at pre-intervention, during the intervention and post intervention. Any disagreements were resolved by group discussion and arbitration.

## Results

Our review found twelve studies, two studies [13,14] were non-evaluable as outcome data from the three aminoglycosides gentamicin, amikacin and tobramycin, were combined (figure 1). Of ten evaluable studies, four were conducted in France, three in Iran, three in Belgium and included 665 participants receiving high-dose amikacin. Four studies reported outcomes following administration of amikacin 30 mg/kg/day as a single dose [15-18], whilst six reported outcomes following administration of amikacin 25 mg/kg/day - four were reports for single doses [19-22] and two for repeated doses [23,24].

**Figure 1.**
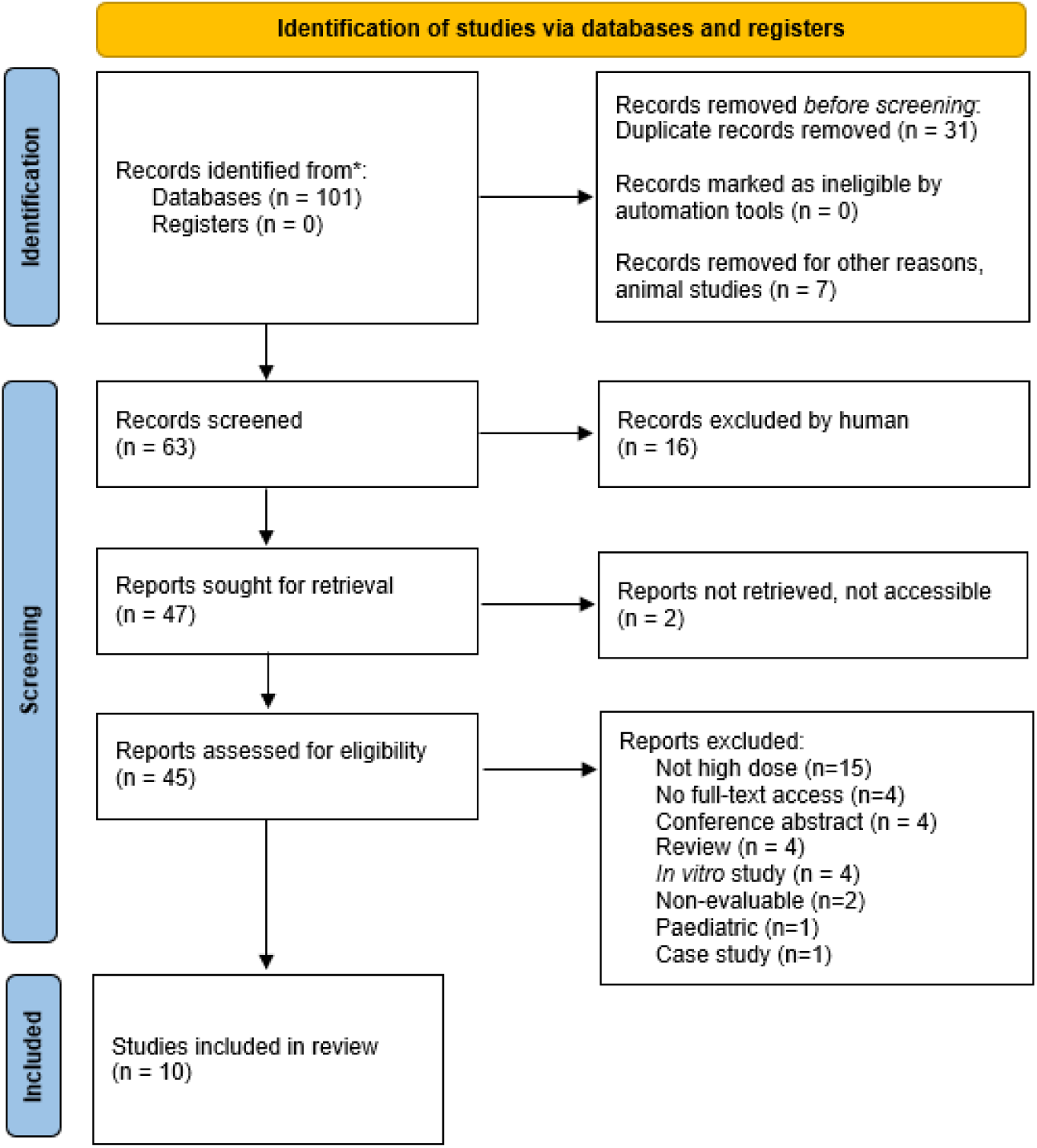
Flow diagram of study selection process. Adapted from PRISMA 2020 Flow Diagram for New Systematic Reviews [11].

### Dosing

Of ten evaluable studies, only two report on repeated doses of amikacin 25 mg/kg/day [23,24] whilst eight report data following a single weight-based dose of amikacin: four at 25 mg/kg [19-22] and four at 30 mg/kg [15-18]. Seven papers adjusted the amikacin dose in extremes of body weight with the characteristics for dosing summarised in Table 1. Najmeddin *et al* do not describe the methodology for dose adjustment in obesity [23], whilst Touchard *et al* capped the dose at 3000 mg (dosing at 25 mg/kg and maximum weight at 120 kg) [21]. The four studies used a Body Mass Index (BMI) of 30 kg/m^2^ as a threshold to commence obese dose adjustment [15-17,24] whilst one using a BMI of 28kg/m^2^ [19]. All studies modifying administered dose in obesity used the following calculation (equation 1) for Adjusted Body Weight (AdjBW) [15-17,19,24].

**Table 1:** Summary of Included and Evaluable Papers

| **Table 1: Summary of Included and Evaluable Papers** |  |  |  |  |  |  |  |  |  |  |
| --- | --- | --- | --- | --- | --- | --- | --- | --- | --- | --- |
| **Author** | **No. Participants** | **Amikacin Dose (mg/kg/day)** | **Adjusted for Obesity** | **Single or Repeated Doses** | **Comparator** | **Indication** | **Exclusions for Renal Impairment** | **Target Peak (mg/ L)** | **Target Trough (mg/ L)** | **Outcomes** |
| *Allou [13]* | 110 | 30 | Yes (BMI > 30kg/m<sup>2</sup>) | First dose only analysed | n/a | Severe sepsis or septic shock | No | 60-80 | < 2.5 | Cmax > 60mg/L in 90/110 participants.<br>Mortality rate lower in pts with Cmax of 60- 80 mg/L compared to a Cmax > 80 mg/L (P = 0.006).<br>Mortality **not** higher in those with Cmax <60 mg/L |
| *Coste [14]* | 89 | 30 | Yes (BMI > 30kg/m<sup>2</sup>) | Single | n/a | Severe sepsis or septic shock | No | > 60 |  | Cmax >60mg/ L in 53 patients (59.6%) after first dose. |
| *Mahmoudi [20]* | 30 | 25 | No | Single | n/a | Severe sepsis | Cr Cl < 60ml/min |  |  | Cmax > 64 µg/ml in 75% of pt. No patient had trough concentration > 5 µg/ml |
| *Najmeddin [21]* | 20,20 | 25 | Yes | Seven days treatment and follow up. | Amikacin 12.5 mg/kg every 12 hours | Sepsis and < 65 years | CrCl < 60ml/min | > 60 |  | Differences of eGFR, rate of AKI and change in serum creatinine were not significant.<br>25mg/kg OD vs 12.5mg/kg BD (Abstract says 12.5mg bd vs 25mg/kg OD but methods state 20mg/kg OD) |
| *Roger [15]* | 47 | 30 | Yes (BMI > 30kg/m<sup>2</sup>) | First dose only analysed | n/a | Severe sepsis in 18-65 years | Requiring renal replacement therapy | > 60 | < 2.5 | Cmax > 60mg/ L was achieved in 36/47 (77%). Second amikacin dose was withheld due to high trough concentrations in 23/47 pts who did not have renal dysfunction |
| *Sadeghi [22]* | 15,18 | 25 | Yes | Seven days treatment, at least 10 days follow up. | Amikacin 15 mg/kg/day | Severe G-ve infection and > 65 years | CrCl < 40ml/min | > 64 | < 5 | 25 mg/kg/day:<br>6/15 pts achieved peaks > 64mg/L whilst 13/15 had troughs > 5mg/L.<br>15mg/kg/day:<br>0/18 pts achieved peaks > 64mg/L whilst 5/18 had troughs > 5mg/ L. |
| *Taccone (2010) [17]* | 74 | 25 | Yes (BMI > 28kg/m<sup>2</sup>) | Single | n/a | Severe sepsis and septic shock | Chronic renal impairment | > 64 | < 5 | Dose adjusted in obesity and low body weight. In 42/74 pts (56.8%) had peaks measured > 64 ml/L whilst 39/74 had troughs >5mg/L. |

| **Table 1: Summary of Included and Evaluable Papers (continued)** |  |  |  |  |  |  |  |  |  |  |
| --- | --- | --- | --- | --- | --- | --- | --- | --- | --- | --- |
| **Author** | **No. Participants** | **Amikacin Dose (mg/kg/day)** | **Adjusted for Obesity** | **Single or Repeated Doses** | **Comparator** | **Indication** | **Exclusions for Renal Impairment** | **Target Peak (mg/ L)** | **Target Trough (mg/ L)** | **Outcomes** |
| *Taccone (2011) [18]* | 13 | 25 | No | Single | n/a | Mixed conditions | Chronic renal impairment | > 64 | < 5 | In 9/13 patients (69%) the peak concentration was >64 mg/L after first dose. |
| *Touchard [19]* | 106 | 25 | Yes | Single | n/a | G –ve infection in ECMO | No | > 60 |  | Cmax was > 60 mg/L in 65/106 patients (61%) after first dose. |
| *Van der Auwera [16]* | 10 | 30 | No | Single | n/a | UTI | Creatinine > 1.5mg/dl |  |  | Concentrations at t=30 mins were approximately double those at t=60 mins. |

**Equation 1**. Adjusted body weight calculation

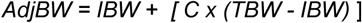

Where *AdjBW* is Adjusted Body Weight (kg), *IBW* is Ideal Body Weight (kg), *TBW* is Total Body Weight (kg), and *C* is the Correction Factor (no units).

Four papers used a correction factor of 0.4 [15,16,19,24] whilst Roger *et al* used 0.43 however the primary source for this correction factor was not cited [17]. Divergence between the papers was observed for the methodology of calculation of IBW, which limits the ability to compare the results (see Supplementary Information – Additional Data)

Only one paper reports dose modifications in low body weight. In patients with a BMI < 20kg/m^2^ Taccone *et al* [19] used the following formula: AdjBW = 1.13 x IBW. In this group, 6/11 (54%) of patients achieved the target of >64 mg/L.

### Pharmacokinetics

Of the eight studies reporting target serum amikacin concentrations, seven listed peak [15-17,19,20,21,23,24] and four listed trough [15,17,19,20] concentrations following amikacin dosing (Table 1). Target peak concentrations (C_max_) were broadly similar at >60 mg/ L [16,17,21,23], >64 mg/ L [19,20,24] and 60-80 mg/ L [15]. From the six studies reporting peak concentrations, between 56.7 and 81.8% of participants achieved target C_max_ following the first amikacin dose.

Five studies reported target trough levels following amikacin dosing, with two papers using <2.5 mg/L [15,17] and three using <5.0 mg/L [19,20,24]. Roger *et al* reported 77% of patients achieving peak levels greater than 60 mg/L following a dose of 30 mg/kg adjusted in obese patients, however in nearly half of these the second dose needed to be withheld due to elevated trough levels (>5 mg/L) [17]. This is supported by Taccone 2010 which reports 52% patients having high trough amikacin levels (<5mg/L) at 24 hours following a dose adjusted in extremes of body weight at 25mg/kg [19].

Two studies reported levels of amikacin measured regularly throughout a 24-hour period following amikacin high-dose administration [18,22]. No studies were identified that used AUC/MIC calculations to measure amikacin treatment outcomes.

### Baseline renal function, nephrotoxicity, audiometry and ototoxicity/vestibular toxicity

Three papers had no exclusions for renal impairment [15,16,21], two papers excluded patients with creatinine clearance (CrCl) <60 mL/min [22,23], and one study excluded patients with CrCl <40 mL/min [24]. Another paper excluded participants requiring renal replacement therapy on critical care [17], while two further papers excluded those with chronic renal failure requiring dialysis [19,20]. Finally, *van der Auwera* excluded patients with a serum creatinine >1.5 mg/dL [18].

Two separate studies from the Iranian research group reported the incidence of acute kidney injury (AKI) in participants receiving high-dose amikacin. One study compared the use of amikacin 12.5 or 25 mg/kg/day for seven days in participants under 65 years [23]. The second reported the use of either 15 or 25 mg/kg/day for seven days in participants age over 65 years; both groups had renal function monitoring for 10 days from the start of treatment (Table 1) [24]. At seven days of follow up, there was no difference in acute kidney injury (AKI), estimated Glomerular Filtration Rate (eGFR) or CrCl deterioration at either dose band in either group. At day 10, there was a statistically significant reduction in CrCl in the over 65 years of age group that was not seen in the under 65 years of age group. Neither auditory nor vestibular toxicities were reported in any of the included studies.

### Assessment of risk of bias

All evaluable non-randomised studies were assessed using ROBINS-I [12], three were considered as critical, three as serious, two as moderate and one as low risk of bias (Figure 2). This was largely influenced by the number of studies assessed with a critical potential for bias due to selection of participants (n=3) and risk of judgement (n=3) and a serious potential of bias due to confounding (n=3) (see and Supplementary Data tables S4, S5, and S6).

**Figure 2:**
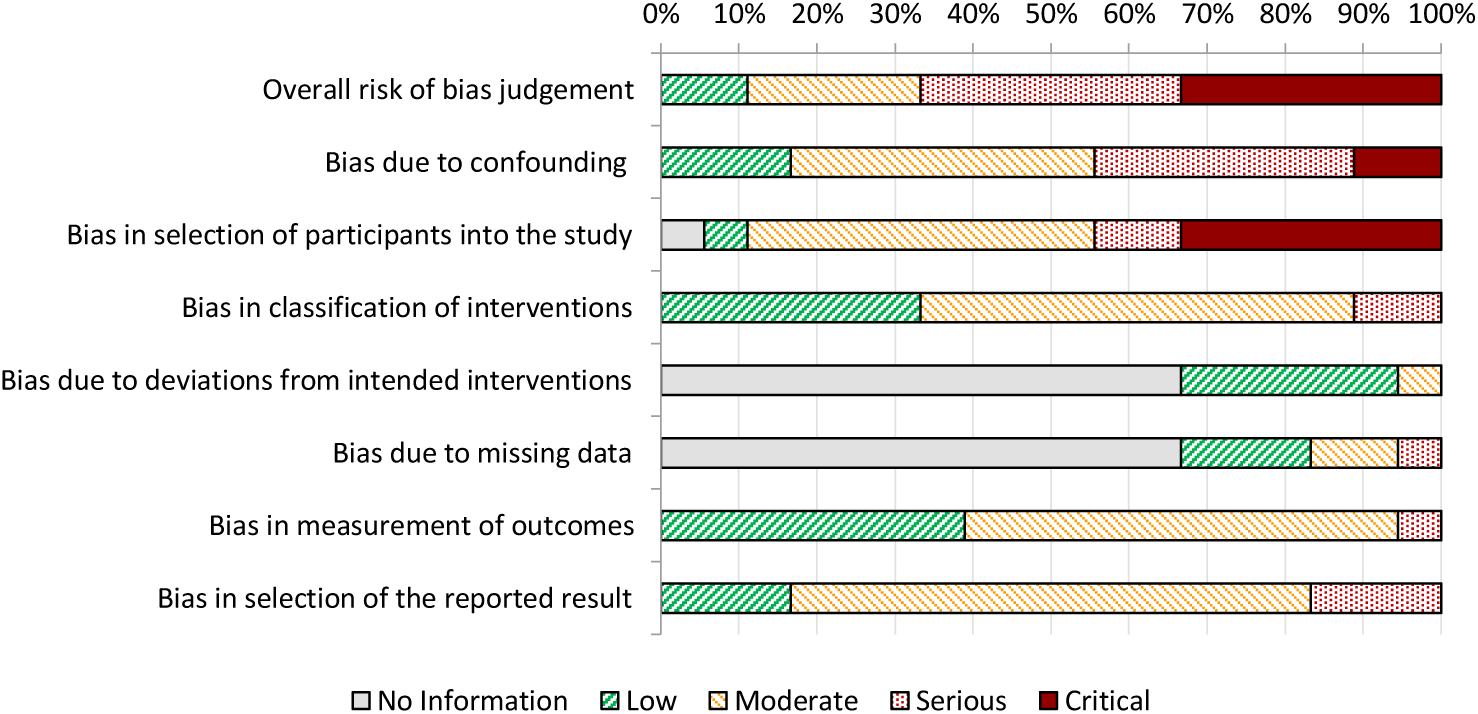
Proportion of reviewed papers falling into low, moderate, serious or critical risk overall (*overall risk of bias judgement*) and for each contributing component of the ROBINS-I tool [12].

## Discussion

Our initial systematic review, and subsequent widening of the included papers, failed to identify high quality evidence to support healthcare professionals in determining the optimal dosing for amikacin – including the most appropriate dosing weight, baseline renal impairment that would preclude high-dose amikacin, the impact on renal and audiological function and the key PK/PD parameters to monitor safety and predict efficacy.

EUCAST guidelines advise that doses above 25 mg/kg are required to obtain sufficient peak concentrations for Gram-negative organisms [6]. Some authors claim that doses between 30-40 mg/kg may be required in critically ill septic patients with increased volume of distribution [13,26,27]. A recent paper by Marsot *et al* reported using doses of up to 40.8 mg/kg TBW in critically unwell patients (n=56) [28].

One ongoing debate with regards to initiating aminoglycosides is which weight to use when calculating doses [29,30] and the literature included in our review does not provide enough evidence to resolve this debate. From the literature, three out of four papers use a BMI >30 kg/m^2^ as the threshold for calculating an AdjBW in dosing calculations in obesity. We found variation in dosing strategies and calculations of IBW which limited our ability to draw valid conclusions from the results of these studies. Taccone et al used an adjusted dosing weight for patients with BMI <20 kg/m2 and found that only 54% of patients achieved a peak amikacin concentration of >64 mg/L; they advised that this patient group might require higher doses than 25 mg/kg of amikacin [19].There is a clear need for further research on the impact of extremes of weight on amikacin pharmacokinetics with an aim to identify optimal dosing strategies across different patient populations.

We were unable to find any evidence that high-dose therapy should be avoided at a different baseline renal function threshold than the BNF standard of creatinine clearance <20 mL/minute or whether courses of high-dose amikacin should have a different maximum course length than the BNF recommended 7 days [31].

There was limited data on the nephrotoxicity of amikacin as most studies were for a single dose where nephrotoxicity was not assessed. Two studies [23,24] looked at repeated dosing of amikacin with durations of treatment of 7 days; interestingly, it was only at 10 days was a difference seen in the creatinine clearance for high-dose amikacin in the older patient cohort in the study by Sadeghi et al [24]. Creatinine clearance is widely used in clinical practice as a surrogate marker for renal function in drugs which are nephrotoxic [32] and while it would seem reassuring that it was only after more than 7 days’ of treatment with high-dose amikacin was any effect seen on renal function, this arm of the study had only 15 patients and so must be interpreted with caution.

None of the included papers conducted audiometric assessment of patients either prior to, during or at the end of amikacin treatment, despite the known ototoxicity of aminoglycosides [33]. Reasons for this would include single dose administration and the large cohort of ITU patients, including critically ill patients with sepsis in which audiometric assessment would not be tenable. As a result, no conclusions on the role of audiometry in high-dose amikacin regimens can be drawn.

Whilst the number of patients that may benefit from the high-dose amikacin regimens may differ depending on local epidemiology, the use of increasing total daily doses of amikacin could be expected to increase risks of toxicity associated with therapy for all exposed. The nephrotoxic and ototoxic complications of aminoglycosides are well documented and associated with excessive dosing and/or supra-therapeutic serum levels [12,34]. The impact of high dosing on vestibular and ototoxicity was not reported in the included papers.

Quantifying the increased risk of high-dose amikacin over that of the traditionally dosed (15 mg/kg/day) amikacin is necessary for clinicians to assess risk/benefit of these two dosing strategies for amikacin. Not all patients will benefit - the majority of Enterobacterales identified within the wild-type distribution have MIC values ≤4 mg/L and high-dose amikacin is likely excessive [5]. The probability that patients will require high-dose amikacin will depend on local epidemiology for Enterobacterales. For *Pseudomonas*, higher MIC values are generally seen and more than 25% of wild-type isolates have MIC value of > 4 mg/L. High-dose amikacin may thus be of more value for pseudomonal infections, although other aminoglycosides such as tobramycin are considered to be more active against Pseudomonas than amikacin [31]. However, it is worth noting that amikacin, and other aminoglycosides, should not be used as monotherapy for non-urinary tract infections, and there are a number of potential alternative agents [5,35].

In the absence of detailed MIC data, or for empiric treatment, EUCAST advise that the higher dosing regimen should be used to ensure coverage across all the wild-type distribution [5]. Yet even with a high-dose regimen, our analysis shows that the optimum PK/PD parameters may not be achieved for almost half of the patients treated with at least 25 mg/kg daily. A proportion receiving high-dose amikacin had raised creatinine levels after 7 days of treatment, suggesting nephrotoxicity, and at the highest end of the dosing range, 30 mg/kg, almost half of the patients treated needed to have a subsequent dose of amikacin withheld due to raised trough levels.

All of the papers included in our review were limited by significant risk of bias, and the signals of higher mortality with high C_max_ could be explained by poor study design and confounding. C_max_ is an unreliable measurement, as it can be affected by method of drug administration and timing of the sample draw [36]. While achievement of an adequate C_max_ is clearly relevant to its antibacterial activity, we would caution against reliance on this parameter based on the low quality of the studies in our review.

## Conclusions

More research to identify the patient population who would most benefit from high-dose amikacin is required. There were only nine papers which were found which could potentially answer this question, and upon systematic review they demonstrated significant divergence due to markedly different dosing regimens utilised. As a result, conclusions are difficult to draw. We propose the development of a consensus treatment guideline in the absence of definitive evidence to support decisions around optimal dosing and monitoring of high-dose amikacin to support healthcare professionals, until further evidence is available.

## Supporting information

Supplementary Information - Literature Review Protocol

## Data Availability

All data produced in the present work are contained in the manuscript

## Transparency declarations

KJF, AJ, CJ, PR, and OT have no declarations of interest to declare. RAH has received educational grants from Pfizer (2021) and attended advisory board for A.Menarini (2022). SH has consulted for Pfizer (2020-21), Eumedica (2020), Kent pharma (2021), Shionogi (2021), BowMed (2021), Gilead (2021).

## Author contributions

CJ conceived the original idea, which then received input from KJF, RAH, SH, PR, OT, and AJ. AJ conducted the searches. All authors contributed equally to assessment of the quality of the studies. AJ extracted the data. All authors contributed equally to the writing and review of the manuscript and approved the final version of the manuscript.

