## Supplementary Information - Literature Review Protocol for "Systematic review of high-dose amikacin regimens for the treatment of Gram-negative infections based on EUCAST dosing recommendations"

**Literature Review Protocol**

**High dose amikacin regimens: how can we maximise the benefits whilst minimising the risks of this dosing strategy in the adult population?**

1.1 **Background**

**1.1.1 Description of the problem**

In January 2020, EUCAST updated their susceptibility breakpoints to reflect increasing resistance and introduced a new classification scheme for determining whether an organism is susceptible, resistant or requires an increased dose of antimicrobial to ensure that key PK/PD targets are achieved.   For amikacin, EUCAST now recommends a dose of 25-30mg/kg once daily for certain resistant organisms to ensure the peak concentration to MIC ratio is achieved.  This dose is significantly higher than traditional dosing regimens of 15mg/kg/day.  EUCAST provided no supporting guidance in the use, administration or monitoring of elevated doses of this potentially toxic drug.

The purpose of this review is to inform consensus guidelines for the dosing, administration and monitoring of patients receiving doses of amikacin ≥20mg/kg/day.

**1.1.2 Description of the intervention**

There is a significant variability in the relationship between the dose of amikacin administered and the plasma level that can be measured in blood, due to factors like renal function and physiological changes that occur in sepsis. As these agents have a narrow therapeutic window, therapeutic drug monitoring (TDM) is considered necessary to ensure the correct dose is used.

**1.1.3 Why it is important to do this review**

This review will cover a frequently monitored agent for which there is a pressing need for clear guidance. In particular to review the scientific basis for both the dosing and TDM of amikacin. From this, a working party will draw up evidence-based guidelines on the use and monitoring of amikacin and provide recommendations that may be adopted into antibiotic policies within individual hospitals.

**1.1.4 Objectives**

To identify high-dose amikacin therapeutic regimes and drug concentrations that are consistent with a good therapeutic outcome and to determine the drug exposures that are related to the adverse events of nephrotoxicity and ototoxicity.

**1.2. Methods**

**1.2.1 Criteria for considering studies for this review**

**1.2.1.1. Types of studies**

A systematic review of amikacin use published in 2016 found one paper that used doses of amikacin >20mg/kg/day.  This search was updated by the present short-life working group in March 2021 and no additional citations were found that met the inclusion criteria.  As a consequence, a new protocol has been developed to specifically search for papers reporting doses of amikacin ≥20 mg/kg/day. This comprehensive review will include observational studies published in full in English.

**1.2.1.2. Types of participants**

Adults treated with amikacin at daily doses at or above 20mg/kg/day.

**1.2.1.3 Types of interventions**

Amikacin ≥20mg/kg/day and therapeutic drug monitoring (TDM).

**1.2.1.4 Types of outcome measures**

***1.2.1.4.1 Primary outcomes***

PK/PD parameters following single or multiple doses of amikacin ≥20mg/kg/day.

TDM results as blood levels reported for cure

TDM results for nephrotoxicity measured by serum creatinine and/or eGFR

Therapeutic cure defined as reduction of fever, improvement in clinical signs, or reduction in inflammatory response.

Adverse events defined as toxicity seen as nephrotoxicity and ototoxicity. Nephrotoxicity to be defined as mild, moderate or severe using the RIFLE criteria

Ototoxicity as a report

**1.2.2 Search methods for identification of studies**

**1.2.2.1 Electronic searches**

Searches will be conducted in Medline, Embase and the Cochrane Central Register of Controlled Trials (CENTRAL), published in *The Cochrane Library.*

The following search strategy will be used by searching in title, abstract and keywords:

#1	Amikacin

#2 	High dose OR Elevated dose OR 20mg OR 21mg OR 22mg OR 23mg OR 24mg OR 24mg OR 26mg OR 27mg OR 28mg OR 29mg OR 30mg OR 31mg OR 32mg OR 33mg OR 34mg OR 35mg OR 2000mg OR 2250mg OR 2500mg OR 2750mg OR 3000mg OR 3250mg OR 3500mg

#3	#1 AND #2

**1.2.2.2 Searching other resources**

Reference lists of included studies will be scanned to seek to identify further studies not identified by electronic searching.

**1.2.3 Data collection and analysis**

**1.2.3.1 Selection of studies**

Studies meeting the inclusion criteria will be identified by two authors (initials) independently and any discrepancies resolved by discussion with other authors. Studies which are excluded after an initial sorting will be recorded with a brief description of the reason for exclusion. Studies will be restricted to English language only.

**1.2.3.2 Data extraction and management**

A data extraction form will be developed to facilitate the collection of data from each included studies. Data extraction will include the following information:

- Lead author and date of publication, dates that the study was conducted
- Participant details including numbers, condition
- Setting and geographical location
- The dose used, frequency of dose and length of treatment
- Numbers of participants with adverse events
- Methods for TDM, including assay and control approaches
- Record of dose or exposure (AUC) and outcome
- Record of dose or exposure (AUC) and toxicity
- Dosing and TDM in participants with adverse events of nephrotoxicity and ototoxicity
- Dosing and TDM in renal impairment or altered pharmacokinetics
- Time of TDM

1.3 **Reporting of Results**

Details of included studies will be recorded in a ‘Characteristics of included studies’ table based on the data extracted. Those studies which were considered but excluded will be listed in an ‘excluded studies’ table together with a brief explanation of the reason for exclusion.
