## Supplementary Information - Additional Data for "Systematic review of high-dose amikacin regimens for the treatment of Gram-negative infections based on EUCAST dosing recommendations"

**Additional Data**

**Amikacin Dose Adjustment in Obesity**

**Equation S1**. Ideal body weight calculation

$$IBW \left( males \right)=50.0+(2.3 \times\left[ H-60 \right])$$

$$IBW \left( females \right)=45.5+(2.3 \times\left[ H-60 \right])$$

Where *IBW* is Ideal Body Weight (kg) and *H* is height (inches).

Variations were observed in the calculations used to determine IBW three studies using the following formula (equation S2) [13, 14, 15].

**Equation S2**. Alternative ideal body weight calculation

$$IBW=Height-100-\frac{\left( Height-150 \right)}{C}+[ 0.4 \times(TBW-IBW)]$$

Where *IBW* is Ideal Body Weight (kg), *TBW* is Total Body Weight (kg), and *C* is the Correction factor (no units, see table S1).

| Table S1. Correction Factor Used in Calculation of Ideal Body Weight (as per Equation S2) | | |
| --- | --- | --- |
|  | Females | Males |
| Allou *et al* [13] | 2 | 4 |
| Coste *et al* [14] | 2 | 4 |
| Roger *et al* [15] | 2.5 | 4 |

| **Table S2.** IBW, AdjBW, and amikacin dosing calculations in obese males using parameters from different studies | | | | | | | | | | | |
| --- | --- | --- | --- | --- | --- | --- | --- | --- | --- | --- | --- |
|  | **Height (cm)** | | | | | | | | | | |
|  | **150** | **155** | **160** | **165** | **170** | **175** | **180** | **185** | **190** | **195** | **200** |
| **Calculated IBW (kg)** | | | | | | | | | | | |
| Allou/ Coste/ Roger | 50 | 53.75 | 57.5 | 61.25 | 65 | 68.75 | 72.5 | 76.25 | 80 | 83.75 | 87.5 |
| Sadeghi | 47.7 | 52.3 | 56.9 | 61.5 | 66.1 | 70.7 | 74.84 | 79.44 | 84.04 | 88.64 | 93.01 |
| **Calculated AdjBW (kg) where ABW is 120kg** | | | | | | | | | | | |
| Allou/ Coste | 78 | 80.25 | 82.5 | 84.75 | 87 | 89.25 | 91.5 | 93.75 | 96 | 98.25 | 100.5 |
| Roger | 80.1 | 82.2375 | 84.375 | 86.5125 | 88.65 | 90.7875 | 92.925 | 95.0625 | 97.2 | 99.3375 | 101.475 |
| Sadeghi | 76.62 | 79.38 | 82.14 | 84.9 | 87.66 | 90.42 | 92.904 | 95.664 | 98.424 | 101.184 | 103.806 |
| *% Difference* | *4.345* | *3.475* | *2.649* | *2.037* | *1.861* | *1.694* | *1.533* | *2.001* | *2.463* | *2.997* | *3.1857* |
| **Amikacin dose (mg)** | | | | | | | | | | | |
| Allou/ Coste (30mg/kg/day) | 2340 | 2407.5 | 2475 | 2542.5 | 2610 | 2677.5 | 2745 | 2812.5 | 2880 | 2947.5 | 3015 |
| Roger  (30mg/kg/day) | 2403 | 2467.13 | 2531.25 | 2595.38 | 2659.5 | 2723.63 | 2787.75 | 2851.88 | 2916 | 2980.13 | 3044.25 |
| Sadeghi  (25mg/kg/day) | 1915.5 | 1984.5 | 2053.5 | 2122.5 | 2191.5 | 2260.5 | 2322.6 | 2391.6 | 2460.6 | 2529.6 | 2595.15 |
| *Difference* | *487.5* | *482.625* | *477.75* | *472.875* | *468* | *463.125* | *465.15* | *460.275* | *455.4* | *450.525* | *449.1* |
| *% Difference* | *20.287* | *19.562* | *18.874* | *18.2199* | *17.5973* | *17.004* | *16.6855* | *16.139* | *15.617* | *15.118* | *14.752* |

| **Table S3.** IBW, AdjBW, and amikacin dosing calculations in obese females using parameters from different studies | | | | | | | | | | | |
| --- | --- | --- | --- | --- | --- | --- | --- | --- | --- | --- | --- |
|  | **Height (cm)** | | | | | | | | | | |
|  | **150** | **155** | **160** | **165** | **170** | **175** | **180** | **185** | **190** | **195** | **200** |
| **Calculated IBW (kg)** | | | | | | | | | | | |
| Allou/ Coste | 50 | 52.5 | 55 | 57.5 | 60 | 62.5 | 65 | 67.5 | 70 | 72.5 | 75 |
| Roger | 50 | 53 | 56 | 59 | 62 | 65 | 68 | 71 | 74 | 77 | 80 |
| Sadeghi | 42.7 | 47.3 | 51.9 | 56.5 | 61.1 | 65.7 | 69.84 | 74.44 | 79.04 | 83.64 | 88.01 |
| **Calculated AdjBW (kg) where ABW is 120kg** | | | | | | | | | | | |
| Allou/ Coste | 78 | 79.5 | 81 | 82.5 | 84 | 85.5 | 87 | 88.5 | 90 | 91.5 | 93 |
| Roger | 80.1 | 81.81 | 83.52 | 85.23 | 86.94 | 88.65 | 90.36 | 92.07 | 93.78 | 95.49 | 97.2 |
| Sadeghi | 73.62 | 76.38 | 79.14 | 81.9 | 84.66 | 87.42 | 89.904 | 92.664 | 95.424 | 98.184 | 100.806 |
| *% Difference* | *8.80196* | *6.6373* | *5.2443* | *3.9071* | *3.3816* | *3.5533* | *3.7185* | *4.4937* | *5.6841* | *6.8076* | *7.7436* |
| **Amikacin dose (mg)** | | | | | | | | | | | |
| Allou/ Coste (30mg/kg/day) | 2340 | 2385 | 2430 | 2475 | 2520 | 2565 | 2610 | 2655 | 2700 | 2745 | 2790 |
| Roger  (30mg/kg/day) | 2403 | 2454.3 | 2505.6 | 2556.9 | 2608.2 | 2659.5 | 2710.8 | 2762.1 | 2813.4 | 2864.7 | 2916 |
| Sadeghi  (25mg/kg/day) | 1840.5 | 1909.5 | 1978.5 | 2047.5 | 2116.5 | 2185.5 | 2247.6 | 2316.6 | 2385.6 | 2454.6 | 2520.15 |
| *Difference* | *562.5* | *544.8* | *527.1* | *509.4* | *491.7* | *474* | *463.2* | *445.5* | *427.8* | *410.1* | *395.85* |
| *% Difference* | *23.4082* | *22.1978* | *21.0368* | *19.9226* | *18.8521* | *17.8229* | *17.0872* | *16.1290* | *15.2058* | *14.3156* | *13.5751* |

**Assessment of Bias**

| **Table S4.** Risk of assessment bias undertaken by Reviewer 1, using the ROBINS-I tool | | | | | | | | |
| --- | --- | --- | --- | --- | --- | --- | --- | --- |
|  | **Pre-intervention** | | **At Intervention** | **Post-intervention** | | | | |
| **Author** | **Bias due to confounding** | **Bias in selection of participants into the study** | **Bias in classification of interventions** | **Bias due to deviations from intended interventions** | **Bias due to missing data** | **Bias in measurement of outcomes** | **Bias in selection of the reported result** | **Risk of bias judgement (overall risk of bias)** |
| Van Der Auwera [16] | Moderate | Critical | Moderate | No Information | No Information | Moderate | Moderate | Critical |
| Taccone [18] | Moderate | Serious | Moderate | No Information | No Information | Moderate | Serious | Serious |
| Mahmoudi [20] | Serious | Critical | Moderate | No Information | No Information | Moderate | Moderate | Critical |
| Najmeddin [21] | Serious | Serious | Serious | No Information | Serious | Moderate | Moderate | Serious |
| Sadeghi [22] | Low | No Information | Low | Low | No Information | Low | Low | Low |
| Roger [15] | Moderate | Moderate | Moderate | Low | Low | Low | Moderate | Moderate |
| Allou [13] | Critical | Critical | Moderate | No Information | No Information | Moderate | Moderate | Critical |
| Touchard [19] | Serious | Moderate | Moderate | No Information | No Information | Moderate | Serious | Serious |
| Coste [14] | Moderate | Moderate | Moderate | Moderate | No Information | Low | Low | Moderate |

| **Table S5.** Risk of assessment bias undertaken by Reviewer 2, using the ROBINS-I tool | | | | | | | | |
| --- | --- | --- | --- | --- | --- | --- | --- | --- |
|  | **Pre-intervention** | | **At Intervention** | **Post-intervention** | | | | |
| **Author** | **Bias due to confounding** | **Bias in selection of participants into the study** | **Bias in classification of interventions** | **Bias due to deviations from intended interventions** | **Bias due to missing data** | **Bias in measurement of outcomes** | **Bias in selection of the reported result** | **Risk of bias judgement (overall risk of bias)** |
| Van Der Auwera [16] | Moderate | Critical | Moderate | No Information | No Information | Moderate | Moderate | Critical |
| Taccone [18] | Moderate | Moderate | Low | Low | Low | Moderate | Serious | Serious |
| Mahmoudi [20] | Serious | Critical | Moderate | No Information | No Information | Moderate | Moderate | Critical |
| Najmeddin [21] | Serious | Moderate | Serious | No Information | No Information | Serious | Moderate | Serious |
| Sadeghi [22] | Low | Moderate | Low | Low | No Information | Low | Moderate | Low |
| Roger [15] | Moderate | Moderate | Low | No Information | Moderate | Low | Moderate | Moderate |
| Allou [13] | Critical | Critical | Moderate | No Information | No Information | Moderate | Moderate | Critical |
| Touchard [19] | Serious | Low | Low | No Information | Low | Low | Moderate | Serious |
| Coste [14] | Low | Moderate | Low | Low | Moderate | Low | Low | Moderate |

| **Table S6**. Comparison of overall risk of bias judgment between Reviewer 1 and Reviewer 2 | | |
| --- | --- | --- |
|  | **Risk of bias judgement (overall risk of bias)** | |
| **Author** | **Reviewer 1** | **Reviewer 2** |
| Van Der Auwera [16] | Critical | Critical |
| Taccone [18] | Serious | Serious |
| Mahmoudi [20] | Critical | Critical |
| Najmeddin [21] | Serious | Serious |
| Sadeghi [22] | Low | Low |
| Roger [15] | Moderate | Moderate |
| Allou [13] | Critical | Critical |
| Touchard [19] | Serious | Serious |
| Coste [14] | Moderate | Moderate |
